# Leveraging Generative AI to Prioritize Drug Repurposing Candidates: Validating Identified Candidates for Alzheimer’s Disease in Real-World Clinical Datasets

**DOI:** 10.1101/2023.07.07.23292388

**Authors:** Chao Yan, Monika E. Grabowska, Alyson L. Dickson, Bingshan Li, Zhexing Wen, Dan M. Roden, C. Michael Stein, Peter J. Embí, Josh F. Peterson, QiPing Feng, Bradley A. Malin, Wei-Qi Wei

## Abstract

Drug repurposing represents an attractive alternative to the costly and time-consuming process of new drug development, particularly for serious, widespread conditions with limited effective treatments, such as Alzheimer’s disease (AD). Emerging generative artificial intelligence (GAI) technologies like ChatGPT offer the promise of expediting the review and summary of scientific knowledge. To examine the feasibility of using GAI for identifying drug repurposing candidates, we iteratively tasked ChatGPT with proposing the twenty most promising drugs for repurposing in AD, and tested the top ten for risk of incident AD in exposed and unexposed individuals over age 65 in two large clinical datasets: 1) Vanderbilt University Medical Center and 2) the *All of Us* Research Program. Among the candidates suggested by ChatGPT, metformin, simvastatin, and losartan were associated with lower AD risk in meta-analysis. These findings suggest GAI technologies can assimilate scientific insights from an extensive Internet-based search space, helping to prioritize drug repurposing candidates and facilitate the treatment of diseases.

## Main

Alzheimer’s disease (AD) is a progressive neurodegenerative disorder that raises major concerns in healthcare due to its irreversibility and high prevalence among older adults^1^. Despite decades of research, treatment options for AD remain limited, leaving patients and families with little hope. Drug repurposing to identify novel therapeutic applications for existing drugs is an attractive additional approach to discovering treatment options compared to the costly and time-consuming process of new drug development alone, particularly for serious, widespread conditions that continue to have few effective treatments, such as AD^2^. In addition to accelerated timelines and lower costs throughout the discovery-to-market process, the approach offers well-established drug safety profiles and expedited clinical translation with enhanced patient accessibility. Nevertheless, the success of drug repurposing hinges on the prompt and accurate identification of promising candidates among a large collection of drugs.

The search for drug repurposing candidates typically relies on a comprehensive review of the scientific literature, focusing on studies that offer evidence of efficacy for certain drugs or their constituent ingredients. Mechanistic insights, preclinical experiments, clinical reports, large-scale observational studies, and drug repurposing databases collectively form the space within which searches are conducted. However, this review process is labor- and time-intensive, requiring researchers to incorporate interdisciplinary expertise in disease mechanisms, molecular biology, pharmacology, clinical research, and bioinformatics. As such, approaches that streamline this process offer an advantage in repurposing efforts.

Recent advancements in generative artificial intelligence (GAI), exemplified by OpenAI’s ChatGPT^3^, have showcased the remarkable capability of AI to understand and respond to diverse inquiries. The comprehension and response capabilities of GAI derive from extensive exposure to a vast corpus from the Internet, nuanced encoding of knowledge, and subsequent optimization of responses that display reasoning processes^4,5^. Beyond answering general questions, GAI has demonstrated effectiveness in specialized medical contexts^6^, including U.S. Medical Licensing Examination queries^7^, clinical decision-making consultations^8,9^, and medical research assessments^10,11^. Notably, ChatGPT is already being leveraged by biotechnology companies to suggest novel pathways for drug targets^12^. However, given its nascent stage and concerns regarding fabrication of information^13,14^, responsible deployment of this tool in the medical setting necessitates comprehensive verification of its functional utility and reliability with clinical data in the real world.

We hypothesized that ChatGPT can function as an AI-driven screening tool to generate drug repurposing candidates for AD. To assess this hypothesis, we provided ChatGPT (model GPT-4) with two sequential prompts. First, we prompted ChatGPT to provide the twenty most promising drug repurposing candidates for AD. Next, we prompted ChatGPT to confirm its previous output and return a final list of drugs (**Fig**. 1a). To account for the probabilistic nature of ChatGPT’s responses, we repeated this process ten times, resulting in a total of 59 unique drug candidates (**Supplementary Table** 1). We confirmed that each candidate appeared in at least one publication discussing their potential use in AD. We then identified the ten most frequently appearing drugs for subsequent testing with clinical data (minimum frequency N=7, maximum frequency N=10).

**Fig. 1:**
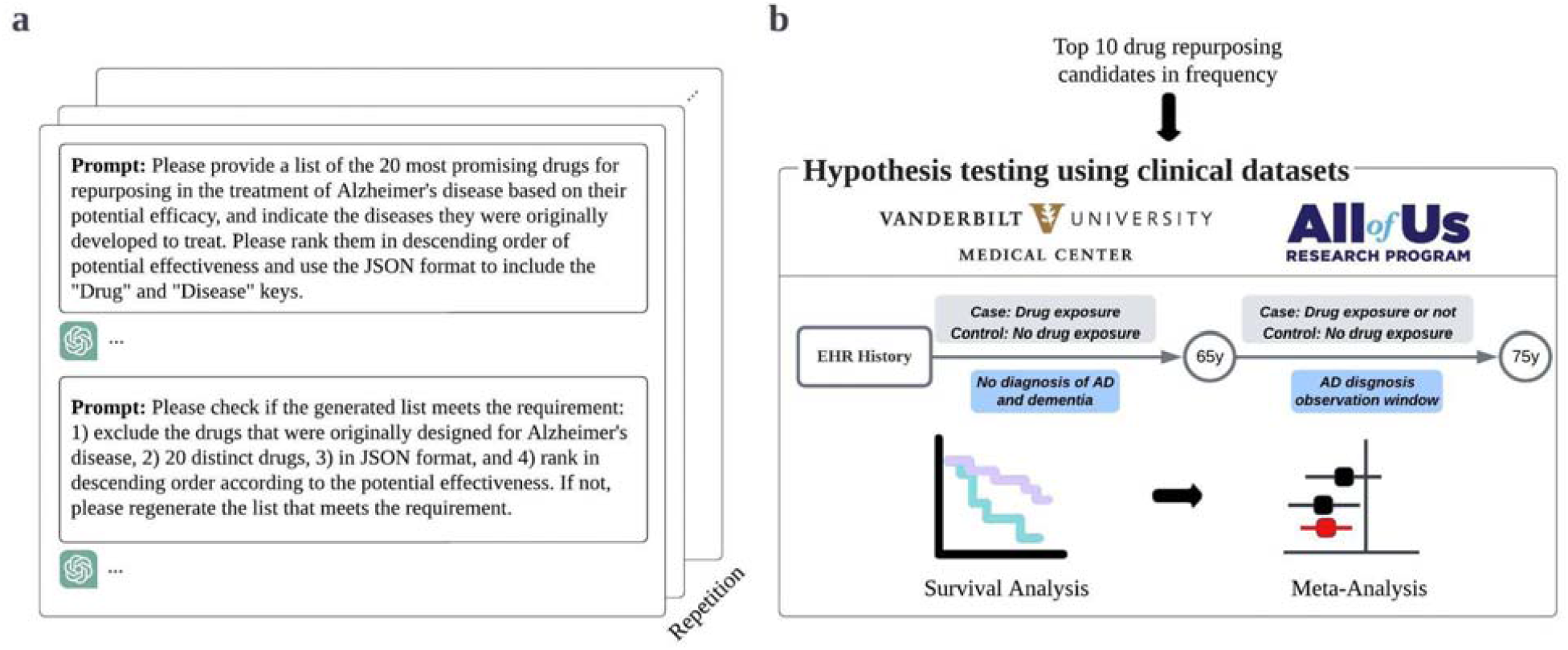
An illustration of the study design. **a**, Employing iterative queries of ChatGPT to recommend twenty drugs for AD repurposing. **b**, Evaluating the potential efficacy of the ten most frequently suggested drugs using electronic health records (EHR) data from two large clinical databases.

**Fig. 2:**
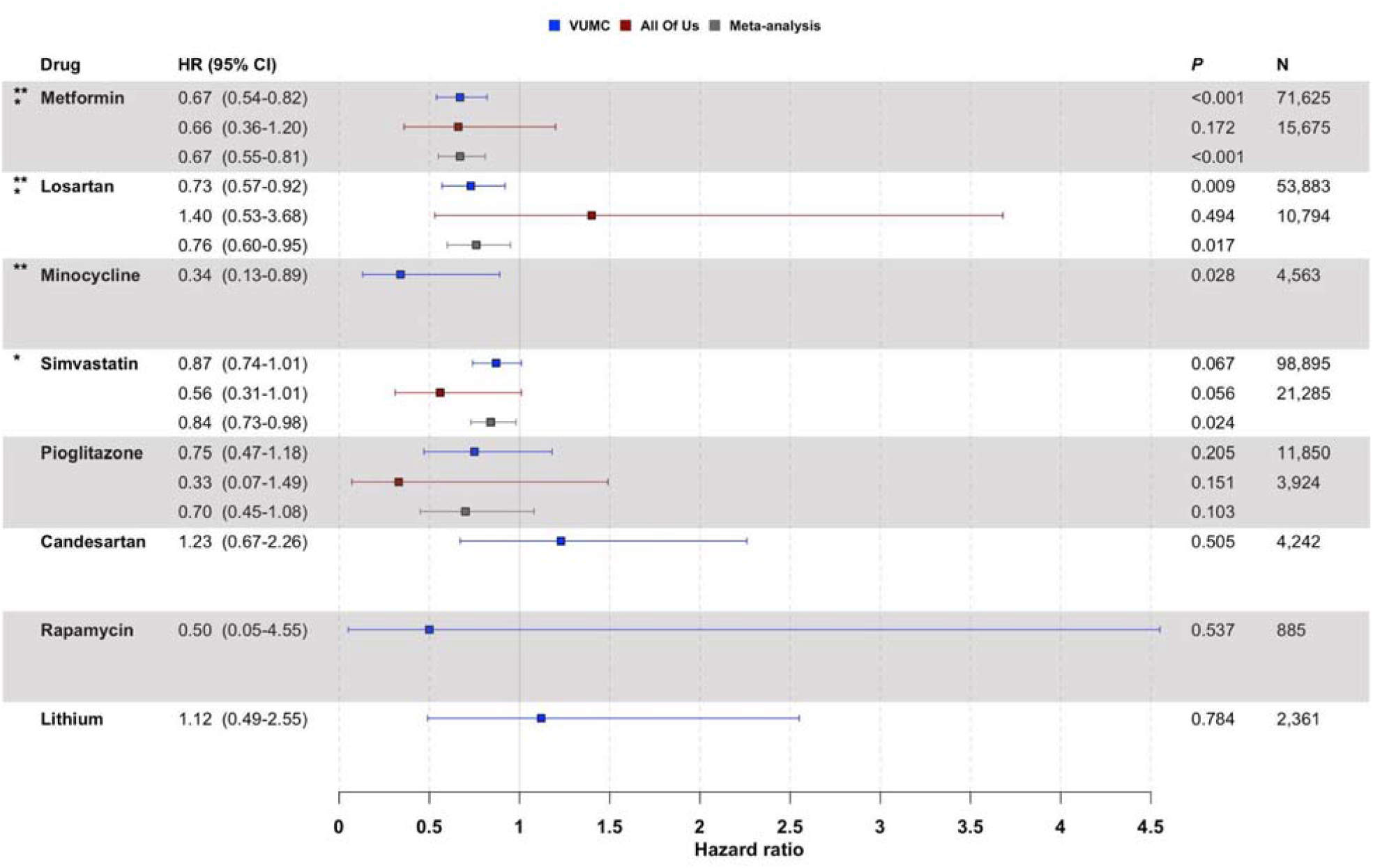
Associations between exposure to ChatGPT-suggested drug repurposing candidates and AD risk. Hazard ratios (HR) and 95% confidence intervals (CI) are shown for VUMC (blue squares), the NIH *All of Us* Research Program (red squares), and the combined meta-analysis (gray squares). ** indicates drugs associated with significantly reduced AD risk using VUMC data (p<0.05); * indicates drugs associated with significantly reduced AD risk in the meta-analysis (p<0.05). To ensure adequate statistical power, we did not report drugs with fewer than five AD cases in the study cohort (i.e., bexarotene and nilotinib in both VUMC and *All of Us*; minocycline, candesartan, rapamycin, and lithium in *All of Us*).

For each generated candidate, we composed two cohorts using de-identified electronic health record (EHR) data from large clinical datasets: 1) Vanderbilt University Medical Center (VUMC), and 2) the National Institutes of Health (NIH) *All of Us* Research Program^15^ (**Fig**. 1b). We employed Cox proportional hazards regression to compare the risk of developing AD between individuals with prior drug exposure and individuals never exposed to the drug. We used age 65 as time zero; prior drug exposure was defined by medication use ≤65 years of age. Each drug-exposed cohort was matched to an unexposed group based on propensity score (PS), using sex, race, EHR length after age 65, and drug-specific comorbidities at age 65 (i.e., at the time of cohort entry) as covariates. Drug-specific comorbidities were selected based on primary clinical indication. Given that the cohort size for a particular drug might not be sufficiently large in the independent datasets, we also performed a meta-analysis to derive a statistically robust estimate of each drug’s hazard ratio.

We observed that three of the top ten ChatGPT recommendations were associated with a significantly reduced risk of AD after ten years of follow-up using VUMC data: the antidiabetic medication metformin (hazard ratio (HR)=0.67, 95% confidence interval (CI): 0.54-0.82, p<1.5?10^−4^), the antihypertensive agent losartan (HR=0.73, 95% CI: 0.57-0.92, p=0.009), and the antibiotic minocycline (HR=0.34, 95% CI: 0.13-0.89, p=0.028) (**Fig**. 2). Though our studies with *All of Us* were limited by smaller sample sizes, metformin showed treatment effects in the expected direction (i.e., HR<1). While not statistically significant at p<0.05, the lipid-lowering medication simvastatin and the antidiabetic medication pioglitazone also exhibited beneficial treatment effects in both the VUMC and *All of Us* data.

In the meta-analysis, we confirmed the protective effect of metformin (HR=0.67, 95% CI: 0.55-0.81, p=6.4×10^−5^). The meta-analysis also revealed a statistically significant protective treatment effect for simvastatin (HR=0.84, 95% CI: 0.73-0.98, p=0.024) that had not been identified in either the VUMC or *All of Us* data in isolation. Losartan was found to have a significant protective treatment effect in meta-analysis as well (HR=0.76, 95% CI: 0.60-0.95, p=0.017); however, the effect estimates from VUMC and *All of Us* were opposing in their directionality.

Inadequate AD case counts (N<5) prevented the evaluation of bexarotene and nilotinib in both VUMC and *All of Us*. The effects of minocycline, candesartan, rapamycin, and lithium could not be tested in *All of Us* for the same reason.

We found that ChatGPT’s utility as a drug repurposing tool resides in its ability to follow instructions pertaining to drug repurposing and rapidly synthesize information from relevant literature. ChatGPT did not propose any FDA-approved drugs for AD, suggesting that it accurately interprets the premise of drug repurposing. In this study, the drugs suggested with the highest frequency by ChatGPT were not novel repurposing candidates for AD, but rather drugs frequently mentioned together with AD in the literature. Antidiabetic drugs such as metformin and pioglitazone have received considerable attention as potential therapeutic candidates for AD, driven by increasing evidence implicating insulin resistance in the pathogenesis of AD^16–18^. Similarly, reported associations between AD and cardiovascular disease have sparked numerous investigations into the repurposing of cardiovascular drugs for AD, including statins and antihypertensive agents such as losartan and candesartan^19–21^. Rapamycin, nilotinib, lithium, and bexarotene have also been heavily explored in AD drug repurposing studies^22–24^.

We observed protective effects against AD for three of the ten drugs most frequently suggested by ChatGPT–metformin, simvastatin, and losartan–in meta-analysis combining data from two large-scale EHRs. Use of metformin, which produced the strongest signal in our meta-analysis, was associated with a 33% decreased risk of incident AD after age 65. Simvastatin and losartan produced more modest effects. In meta-analysis, simvastatin was associated with a 16% decreased risk of AD, while losartan was associated with a 24% decreased risk of AD. Whereas metformin and simvastatin were found to have consistent treatment effects (HR<1) in both VUMC and *All of Us*, losartan had conflicting treatment effects (statistically significant HR<1 using VUMC data, non-significant HR>1 using *All of Us* data). This suggests that losartan’s protective treatment effect in meta-analysis may have been driven by the larger sample size from VUMC. Despite supporting findings for these three drugs in previous studies, much remains unknown about the mechanisms by which these drugs affect AD pathophysiology and pathology, and population-based studies have not provided conclusive results^25–27^. Further investigation in preclinical and clinical studies will be needed to ascertain the viability of these drugs in decreasing risk of AD.

Our findings suggest that ChatGPT can generate quality hypotheses for drug repurposing. ChatGPT expedites the process of extensive literature review, which has become infeasible for humans to perform alone. With minimal costs, ChatGPT has the capacity and scalability to substantially accelerate the review process, allowing researchers to focus on testing and validating the hypotheses. Moreover, the anticipated regular updates of ChatGPT (which provide access to new Internet content) and its search engine plugins allow for consistently up-to-date and uninterrupted drug repurposing research. Furthermore, combining ChatGPT-powered hypotheses with robust verification using real-world clinical datasets provides a cost-effective pipeline to investigate preliminary signals before allocating additional resources to extensive research and clinical trials. This validation process serves as a critical balancing force to disprove invalid hypotheses, assuaging concerns about adverse consequences of AI hallucinations–a major criticism of ChatGPT use. Despite these advantages, any pipelines incorporating ChatGPT must account for the possibility of overlooked, but promising, repurposing candidates, which can transpire when candidates exhibit low occurrence in the literature or necessitate complex reasoning ability based on indirect evidence that surpasses ChatGPT’s capabilities.

Our study has several limitations of note. First, we relied upon frequency to prioritize drug candidates; however, the number of times a repurposing candidate appears in ChatGPT queries may not be directly related to its promise in treating disease. Second, EHRs can contain missing or incomplete data^28^, and discontinuities in medication adherence may not be reported with perfect fidelity, creating possibilities for misclassification of outcome or exposure. Third, despite the use of two large EHRs, we still did not have adequate statistical power for hypothesis testing of less common drugs (e.g., nilotinib). Fourth, while our study evaluated drug exposure broadly as any-time, any-dose exposure ≤65 years of age, there exist many opportunities for deeper phenotyping in characterizing drug exposure. Fifth, we sought to control for a single primary indication for each drug using MEDI; however, we were unable to establish a clear primary indication for several drugs (i.e., nilotinib, bexarotene, minocycline, and rapamycin).

Furthermore, a fully balanced covariate distribution was not achieved for metformin and simvastatin (standardized mean difference >0.1 for EHR length after 65 and drug-specific comorbidities), suggesting there may be some residual confounding (although likely to bias towards the null). Sixth, this study cannot establish causal effects or mechanisms as might be the case in a clinical trial. Lastly, although ChatGPT exhibits exceptional response quality for general queries, further research is required to benchmark a range of GAI models and their fine-tuned variants for greatest effectiveness and reliability in supporting biomedical tasks, particularly drug repurposing.

Still, this proof-of-concept study showcases the feasibility of employing ChatGPT as an AI-driven hypothesis generator for drug repurposing, enabling the prompt generation of a promising list of drugs for subsequent testing in EHRs, using AD as a case study. Our findings suggest that ChatGPT is able to encode valuable insights concerning novel potential therapeutic utilities for existing drugs by comprehensively synthesizing literature, and can subsequently decode this knowledge when responding to queries. Pipelines that leverage the capabilities of ChatGPT offer a streamlined new framework for drug repurposing that can be applied to numerous diseases.

## Methods

Usage of *All of Us* data was approved by the NIH *All of Us* Research Program. All EHR data from VUMC was de-identified, such that this study was deemed to be exempt by the Institutional Review Board.

### Interactions with ChatGPT

In this study, we interacted with OpenAI’s ChatGPT (GPT-4) to generate promising drug repurposing candidates for AD. These interactions were conducted in May 2023, at which time the technology had access to information accumulated until September 2021. Ten independent queries were performed, ensuring that each query did not serve as the context for another. Each query consisted of two prompts. The first prompt described the instructions for generating drug repurposing candidates, whereas the second prompt asked ChatGPT to self-correct its output from the previous prompt.

- **Prompt 1:** Please provide a list of the 20 most promising drugs for repurposing in the treatment of Alzheimer’s disease based on their potential efficacy, and indicate the diseases they were originally developed to treat. Please rank them in descending order of potential effectiveness and use the JSON format to include the “ Drug” and “ Disease” keys.
- **Prompt 2:** Please check if the generated list meets the requirement: 1) exclude the drugs that were originally designed for Alzheimer’s disease, 2) 20 distinct drugs, 3) in JSON format, 4) rank in descending order according to the potential effectiveness. If not, please regenerate the list that meets the requirement.

In our queries, we intentionally emphasized drugs’ original purposes to encourage ChatGPT to distinguish between the drugs originally intended to treat AD and those used in treating other diseases. This helped to limit the possibility that candidates with original use in AD were returned in the final list of each query. We also imposed a specific format for the drugs returned in the queries to facilitate subsequent processing. We also asked ChatGPT to rank drugs according to their potential effectiveness. While ChatGPT claimed that it “ cannot rank the generated drugs with respect to their potential effectiveness since the data is not definitive and is constantly evolving” in multiple responses, we sought to emphasize the notion of effectiveness during the drug generation process. It is important to note that we did not use the order of drugs in the generated lists for subsequent drug selection.

### Data source

We performed our clinical validation studies using de-identified EHR data from 1) Vanderbilt University Medical Center (VUMC), a major academic medical center in Nashville, Tennessee, and 2) the *All of Us* Research Program run by the National Institutes of Health (NIH), a U.S. nation-wide clinical database. VUMC’s de-identified EHR database contains longitudinal clinical data including diagnosis codes, lab values, and medications for over three million patient records^29^. The NIH *All of Us* Research Program database contained de-identified EHR data for over 235,000 participants at the time of this study^15^. The EHR data in both resources is standardized according to the Observational Medical Outcomes Partnership (OMOP) Common Data Model^30^, allowing for reproducible cohort formation and characterization of drug exposures in the two databases.

### Study cohort

For each candidate drug, we conducted a retrospective cohort study using age 65 as time 0. Each study was limited to individuals aged 65 or older with no prior diagnosis of AD. We excluded individuals with a diagnosis of non-Alzheimer’s dementia (vascular dementia, diffuse Lewy body disease, frontotemporal dementia, mixed dementia, and dementia associated with Parkinson’s disease), individuals without EHR follow-up after age 65, and individuals with missing demographic characteristics.

We defined a confirmed diagnosis of AD as patients with at least one AD diagnosis code in their EHR using ICD-9-CM code 331.0 and ICD-10-CM codes G30.1, G30.8, and G30.9. We have previously shown that using ICD codes to phenotype AD patients has a high PPV (94%) in VUMC’s de-identified EHR database^31^.

To capture all relevant drug exposures when creating the drug-exposed group, medications were mapped to their ingredients using RxNorm^32^. Individuals with at least one recorded exposure to the drug of interest occurring at ≤65 years of age were considered to be exposed. Individuals whose first record of drug exposure occurred after age 65 were excluded from the analysis.

We gathered demographic characteristics (sex and race), remaining chart length, and comorbidities at age 65 to generate a propensity score (PS) for matching. The comorbidities were selected to mitigate potential confounding by indication. We used MEDI^33^, an ensemble medication indication resource, to identify the primary clinical indication for each drug repurposing candidate. MEDI contains over 63,000 medication-indication pairs with indication prevalence evaluated using EHR data. We queried MEDI for the highest prevalence indications for each drug and used these to define a single primary indication for the drug. If there was no consensus among the top indications, a primary indication for the drug was not defined. MEDI reports medication indications using only ICD-9-CM; as such, we mapped the ICD-9-CM code(s) comprising the primary indications to ICD-10-CM codes using the General Equivalence Mappings developed by the Centers for Medicare & Medicaid Services. **Supplementary Table** 2 reports the set of ICD-9-CM and ICD-10-CM codes relied upon to define the comorbidities and the drugs they pertain to. A confirmed comorbidity status was defined as disease diagnosed at the start of follow-up (i.e., at ≤65 years of age).

We applied 2:1 PS matching (nearest-neighbor algorithm, caliper = 0.1) with sex, race, length of EHR after age 65, and relevant drug-specific comorbidities as covariates to form comparable drug-exposed and unexposed cohorts for each suggested drug repurposing candidate. PS matching was performed using the MatchIt R package^34^. The participant counts for each drug after matching (AD/exposed, no AD/exposed, AD/unexposed, and no AD/unexposed) are provided in **Supplementary Table** 3. The covariate balance between the drug-exposed and unexposed groups after matching is provided in **Supplementary Table** 4.

Based on our study design, an individual with a history of exposure to multiple drug repurposing candidates could be included in more than one drug-exposed cohort. We did not consider potential compound effects resulting from multiple drug exposures.

### Statistical analysis

All survival analyses were performed using Cox proportional hazards regression models. Each model compared the risk of AD in individuals exposed to a drug repurposing candidate and PS-matched individuals never exposed to the drug. Follow-up ended at the first of 1) AD diagnosis, last recorded EHR observation, or 3) ten years. We censored observations after ten years of EHR follow-up (i.e., at age 75) to minimize differential loss to follow-up. To ensure adequate statistical power, we did not report drugs with fewer than five AD cases included in the final study cohort. We used p<0.05 as our significance threshold given the small number of tests (N=10).

Meta-analysis of hazard ratios was performed using NCSS statistical software^35^. Cochran’s Q test was used to assess heterogeneity. Meta-analysis was performed under a fixed-effects model.

## Data Availability

The VUMC dataset used in this study is available upon request from the corresponding authors and subsequent institutional approval. The All of Us dataset can be accessed through the Researcher Workbench by following the detailed data application process outlined at https://www.researchallofus.org.

## Data Availability

The VUMC dataset used in this study is available upon request from the corresponding authors and subsequent institutional approval. The *All of Us* dataset can be accessed through the Researcher Workbench by following the detailed data application process outlined at http://www.researchallofus.org/.

## Code Availability

The source code associated with this study is publicly available at: https://github.com/monikagrabowska/GPT4_AD_Drug_Repurposing.

## Acknowledgements

This study was supported by the National Institute of General Medical Sciences of the National Institutes of Health under award numbers R01GM139891, R35GM131770 and the National Institute of Aging of the National Institutes of Health under award numbers R01AG069900, F30AG080885.

## Author Information

C.Y. and M.E.G. contributed equally and share the first authorship. W.Q.W. and B.A.M. jointly supervised this research and share the senior authorship. W.Q.W., C.Y., and M.E.G. conceived and designed this study. M.E.G and C.Y. performed the data collection, curation, and experiments and analyzed the results. Q.P.F., A.L.D. and C.M.S. provided guidance on cohort selection and survival analysis study design. B.L., Z.W., D.M.R., P.J.E., and J.F.P. critically reviewed the paper and contributed important intellectual content. C.Y. and M.E.G. wrote the original draft. A.L.D., M.E.G., and C.Y. led paper revision. All authors approved this study.

## References

1. Matthews, K. A. et al. Racial and ethnic estimates of Alzheimer’s disease and related dementias in the United States (2015-2060) in adults aged ≥65 years. Alzheimers. Dement. 15, 17–24 (2019).

2. Pushpakom, S. et al. Drug repurposing: progress, challenges and recommendations. Nat. Rev. Drug Discov. 18, 41–58 (2019).

3. OpenAI. Introducing ChatGPT. November 30, 2022 (https://openai.com/blog/chatgpt).

4. Singhal, K. et al. Large language models encode clinical knowledge. arXiv [cs.CL] (2022).

5. Liu, H. et al. Evaluating the logical reasoning ability of ChatGPT and GPT-4. arXiv [cs.CL] (2023).

6. Lee, P. et al. Benefits, limits, and risks of GPT-4 as an AI chatbot for medicine. N. Engl. J. Med. 388, 1233–1239 (2023).

7. Kung, T. H. et al. Performance of ChatGPT on USMLE: Potential for AI-assisted medical education using large language models. PLOS Digit. Health 2, e0000198 (2023).

8. Ayers, J. W. et al. Comparing physician and artificial intelligence chatbot responses to patient questions posted to a public social media forum. JAMA Intern. Med. (2023) doi:10.1001/jamainternmed.2023.1838.

9. Liu, S. et al. Using AI-generated suggestions from ChatGPT to optimize clinical decision support. J. Am. Med. Inform. Assoc. (2023) doi:10.1093/jamia/ocad072.

10. Cahan, P. & Treutlein, B. A conversation with ChatGPT on the role of computational systems biology in stem cell research. Stem Cell Reports 18, 1–2 (2023).

11. Aydın, Ö. & Karaarslan, E. OpenAI ChatGPT generated literature review: Digital twin in healthcare. SSRN Electron. J. (2022) doi:10.2139/ssrn.4308687.

12. Savage, N. Drug discovery companies are customizing ChatGPT: here’s how. Nat. Biotechnol. 41, 585–586 (2023).

13. Májovský, M. et al. Artificial intelligence can generate fraudulent but authentic-looking scientific medical articles: Pandora’s box has been opened. J. Med. Internet Res. 25, e46924 (2023).

14. Kung, T. H. et al. Performance of ChatGPT on USMLE: Potential for AI-assisted medical education using large language models. PLOS Digit. Health 2, e0000198 (2023).

15. All of Us Research Program Investigators et al. The “ All of Us” Research Program. N. Engl. J. Med. 381, 668–676 (2019).

16. Kellar, D. & Craft, S. Brain insulin resistance in Alzheimer’s disease and related disorders: mechanisms and therapeutic approaches. Lancet Neurol. 19, 758–766 (2020).

17. Leclerc, M. et al. Cerebrovascular insulin receptors are defective in Alzheimer’s disease. Brain 146, 75–90 (2023).

18. Michailidis, M. et al. Antidiabetic drugs in the treatment of Alzheimer’s disease. Int. J. Mol. Sci. 23, 4641 (2022).

19. Leszek, J. et al. The links between cardiovascular diseases and Alzheimer’s disease. Curr. Neuropharmacol. 19, 152–169 (2021).

20. Torrandell-Haro, G. et al. Statin therapy and risk of Alzheimer’s and age-related neurodegenerative diseases. Alzheimers Dement. (N. Y.) 6, e12108 (2020).

21. Adesuyan, M. et al. Antihypertensive agents and incident Alzheimer’s disease: A systematic review and meta-analysis of observational studies. J. Prev. Alzheimers Dis. 9, 715–724 (2022).

22. Kaeberlein, M. & Galvan, V. Rapamycin and Alzheimer’s disease: Time for a clinical trial? Sci. Transl. Med. 11, eaar4289 (2019).

23. Nobili, A. et al. Nilotinib: from animal-based studies to clinical investigation in Alzheimer’s disease patients. Neural Regen. Res. 18, 803–804 (2023).

24. Tousi, B. The emerging role of bexarotene in the treatment of Alzheimer’s disease: current evidence. Neuropsychiatr. Dis. Treat. 11, 311–315 (2015).

25. Ha, J. et al. Association of metformin use with Alzheimer’s disease in patients with newly diagnosed type 2 diabetes: a population-based nested case-control study. Sci. Rep. 11, 24069 (2021).

26. Jeong, S.-M. et al. Association between statin use and Alzheimer’s disease with dose response relationship. Sci. Rep. 11, 15280 (2021).

27. Kehoe, P. G. et al. Safety and efficacy of losartan for the reduction of brain atrophy in clinically diagnosed Alzheimer’s disease (the RADAR trial): a double-blind, randomised, placebo-controlled, phase 2 trial. Lancet Neurol. 20, 895–906 (2021).

28. Haneuse, S. et al. Assessing missing data assumptions in EHR-based studies: A complex and underappreciated task. JAMA Netw. Open 4, e210184 (2021).

29. Zheng, N. S. et al. A retrospective approach to evaluating potential adverse outcomes associated with delay of procedures for cardiovascular and cancer-related diagnoses in the context of COVID-19. J. Biomed. Inform. 113, 103657 (2021).

30. Data standardization – OHDSI. Ohdsi.org https://www.ohdsi.org/data-standardization/.

31. Thakkar, R. et al. Developing a universal phenotyping algorithm to identify patients with clinically diagnosed and probable Alzheimer’s disease using electronic health record data. Alzheimers. Dement. 18, (2022).

32. Nelson, S. J. et al. Normalized names for clinical drugs: RxNorm at 6 years. J. Am. Med. Inform. Assoc. 18, 441–448 (2011).

33. Wei, W.-Q. et al. Development and evaluation of an ensemble resource linking medications to their indications. J. Am. Med. Inform. Assoc. 20, 954–961 (2013).

34. Ho, D. E. et al. MatchIt: Nonparametric Preprocessing for Parametric Causal Inference. J. Stat. Softw. 42, (2011).

35. Penman, N. & Pastore, F. G. Statistical software. Ncss.com http://ncss.com/software/ncss (2012).

