## Supplementary for "Leveraging Generative AI to Prioritize Drug Repurposing Candidates: Validating Identified Candidates for Alzheimer’s Disease in Real-World Clinical Datasets"

**Supplementary Table 1. The full list of drug repurposing candidates generated by ChatGPT (N=59).** Drug frequencies observed across ten independent queries are indicated. Exemplary references are provided by the authors to verify the candidate's demonstrated potential for Alzheimer's disease (AD).

| **Drug candidate** | **Original purpose generated by ChatGPT** | **Frequency** | **Examples of references** |
| --- | --- | --- | --- |
| Simvastatin | Hypercholesterolemia | 10 | 1,2 |
| Metformin | Type 2 diabetes | 10 | 3,4 |
| Losartan | Hypertension | 10 | 5,6 |
| Pioglitazone | Type 2 Diabetes | 10 | 7,8 |
| Candesartan | Hypertension | 9 | 9.10 |
| Rapamycin | Organ transplant rejection | 9 | 11,12 |
| Bexarotene | Cutaneous T-cell lymphoma | 8 | 13,14 |
| Lithium | Bipolar disorder | 7 | 15,16 |
| Minocycline | Bacterial infections | 7 | 17,18 |
| Nilotinib | Chronic Myeloid Leukemia | 7 | 19,20 |
| Montelukast | Asthma and allergies | 6 | 21,22 |
| Saracatinib | Cancer (solid tumors) | 6 | 23,24 |
| Fluoxetine | Depression | 6 | 25,26 |
| Carvedilol | Congestive heart failure, hypertension | 6 | 27,28 |
| Riluzole | Amyotrophic lateral sclerosis | 5 | 29,30 |
| Citalopram | Depression | 5 | 31,32 |
| Doxycycline | Bacterial infections | 5 | 33,34 |
| Atorvastatin | Hypercholesterolemia | 4 | 35,36 |
| Masitinib | Cancer (gastrointestinal stromal tumors) | 4 | 37,38 |
| Nilvadipine | Hypertension | 3 | 39,40 |
| Mebendazole | Parasitic Infections | 3 | 41 |
| Levetiracetam | Epilepsy | 3 | 42,43 |
| Ibudilast | Asthma | 3 | 44,45 |
| Methylene blue | Malaria | 3 | 46,47 |
| Latrepirdine | Allergies | 3 | 48,49 |
| Valproate | Epilepsy | 2 | 50,51 |
| Trazodone | Depression | 2 | 52,53 |
| Nortriptyline | Depression | 2 | 54 |
| Methotrexate | Rheumatoid Arthritis | 2 | 55,56 |
| J147 | Neurological dysfunction | 2 | 57,58 |
| Rosiglitazone | Type 2 Diabetes | 2 | 59,60 |
| Telmisartan | Hypertension | 2 | 61,62 |
| Exenatide | Type 2 diabetes | 2 | 63,64 |
| Etanercept | Rheumatoid Arthritis | 2 | 65,66 |
| Dextromethorphan | Cough | 2 | 67,68 |
| Valproic acid | Epilepsy | 1 | 69,70 |
| Valsartan | Hypertension | 1 | 71,72 |
| Sodium valproate | Epilepsy | 1 | 50 |
| Tolcapone | Parkinson's disease | 1 | 73 |
| Rivaroxaban | Deep vein thrombosis and pulmonary embolism | 1 | 74 |
| Nebivolol | Hypertension | 1 | 75 |
| Sildenafil | Erectile dysfunction | 1 | 76 |
| Acamprosate | Alcohol dependence | 1 | 77 |
| Naproxen | Inflammation | 1 | 78 |
| Colchicine | Gout | 1 | 79 |
| Amiloride | Hypertension | 1 | 80 |
| Amitriptyline | Depression and Neuropathic Pain | 1 | 81 |
| Azeliragon | Type 2 Diabetes* | 1 | 82 |
| Baclofen | Muscle Spasticity | 1 | 83 |
| Cannabidiol | Epilepsy | 1 | 84 |
| Cimetidine | Peptic Ulcers | 1 | 85 |
| Clemastine | Allergy, Hay fever | 1 | 86 |
| Dexamethasone | Inflammation | 1 | 87 |
| Naltrexone | Opioid and alcohol dependence | 1 | 88 |
| Exendin-4 | Type 2 diabetes | 1 | 89 |
| Ganaxolone | Epilepsy | 1 | 90 |
| Hydroxychloroquine | Malaria, Rheumatoid Arthritis, Lupus | 1 | 91 |
| Ibuprofen | Pain and Inflammation | 1 | 92 |
| Intranasal insulin | Diabetes | 1 | 93 |

*****Azeliragon was first tested for use in AD by Pfizer, but has been proposed for treating a variety of diseases, including type 2 diabetes, glioblastoma, and cancer.

**Supplementary Table 2. International Classification of Diseases (ICD-9-CM and ICD-10-CM) codes used to define drug-specific comorbidities based on primary clinical indication.**

| **Drug** | **Clinical indication** | **Diagnosis codes** |
| --- | --- | --- |
| Lithium | Bipolar or depressive disorder | **ICD-9-CM:** 296.80, 311  **ICD-10-CM:** F31.9, F32.9 |
| Metformin, Pioglitazone | Type 2 diabetes | **ICD-9-CM:** 250.00  **ICD-10-CM:** E11.9 |
| Simvastatin | Hyperlipidemia | **ICD-9-CM:** 272.0, 272.2, 272.4, 413.9, 414.00, 786.50  **ICD-10-CM:** E78.2, E78.5, I20.8, I20.9, I25.10, R07.9 |
| Candesartan, Losartan | Hypertension | **ICD-9-CM:** 401.1, 401.9  **ICD-10-CM:** I10 |
| Rapamycin | N/A | N/A |
| Nilotinib | N/A | N/A |
| Minocycline | N/A | N/A |
| Bexarotene | N/A | N/A |

**Supplementary Table 3. Alzheimer's disease (AD) event counts stratified by drug exposure status for the ten drug repurposing candidates most frequently recommended by ChatGPT.** In accordance with the *All of Us* Data and Statistics Dissemination Policy, participant counts between 1 and 20 are not reported directly.

|  | **VUMC**  (N) | | | | **All of Us**  (N) | | | |
| --- | --- | --- | --- | --- | --- | --- | --- | --- |
|  | **Exposed** | | **Unexposed** | | **Exposed** | | **Unexposed** | |
| **Drug** | **AD** | **No AD** | **AD** | **No AD** | **AD** | **No AD** | **AD** | **No AD** |
| Metformin | 145 | 23,730 | 217 | 47,533 | ≤20 | 5,209 | 28 | 10,422 |
| Losartan | 93 | 17,868 | 255 | 35,667 | ≤20 | 3,591 | ≤20 | 7,186 |
| Minocycline | 5 | 1,516 | 29 | 3,013 | 0 | 396 | ≤20 | 790 |
| Simvastatin | 260 | 32,705 | 458 | 65,472 | ≤20 | 7,081 | 48 | 14,142 |
| Pioglitazone | 25 | 3,925 | 67 | 7,833 | ≤20 | 1,306 | ≤20 | 2,604 |
| Candesartan | 16 | 1,398 | 26 | 2,802 | 0 | 144 | 0 | 288 |
| Rapamycin | 1 | 294 | 4 | 586 | 0 | 60 | 0 | 120 |
| Lithium | 9 | 778 | 16 | 1,558 | 0 | 182 | ≤20 | 362 |
| Bexarotene | 0 | 108 | 3 | 213 | 0 | ≤20 | 0 | ≤20 |
| Nilotinib | 0 | 33 | 1 | 65 | 0 | ≤20 | 0 | ≤20 |

**Supplementary Table 4. Standardized mean differences for covariates after propensity score matching.**

|  | **VUMC** | | | | **All of Us** | | | |
| --- | --- | --- | --- | --- | --- | --- | --- | --- |
| **Drug** | **Sex** | **Race** | **EHR length after 65** | **Comorbidity** | **Sex** | **Race** | **EHR length after 65** | **Comorbidity** |
| Metformin | 0.0161 | -0.0342 | 0.5734 | 1.0143 | -0.0261 | -0.0472 | 0.4973 | 1.2600 |
| Losartan | 0.0030 | -0.0014 | 0.0002 | 0.0002 | -0.0102 | 0.0031 | 0.0002 | 0.0000 |
| Minocycline | 0 | 0 | 0 | N/A | 0 | 0 | 0 | N/A |
| Simvastatin | -0.0125 | -0.2056 | 0.2072 | 0.1983 | -0.0033 | -0.0170 | 0.0357 | 0.0000 |
| Pioglitazone | 0.0000 | 0.0023 | 0.0000 | 0.0000 | -0.0287 | 0.0040 | 0.0019 | -0.0016 |
| Candesartan | 0 | 0 | 0 | 0 | -0.0242 | -0.0004 | 0.0078 | 0.0000 |
| Rapamycin | 0 | 0 | 0 | N/A | 0.0170 | 0.0010 | -0.0032 | N/A |
| Lithium | -0.0026 | -0.0062 | 0.0013 | 0.0000 | 0 | 0 | 0 | 0 |
| Bexarotene | 0 | 0 | 0 | N/A | 0 | 0 | 0 | N/A |
| Nilotinib | 0 | 0 | 0 | N/A | 0 | 0 | 0 | N/A |

**References**

1. Serrano-Pozo, A. *et al.* Effects of simvastatin on cholesterol metabolism and Alzheimer disease biomarkers. *Alzheimer Dis. Assoc. Disord.* 24, 220–226 (2010).
2. Langness, V. F. *et al.* Cholesterol-lowering drugs reduce APP processing to Aβ by inducing APP dimerization. *Mol. Biol. Cell* 32, 247–259 (2021).
3. Liao, W. *et al.* Deciphering the roles of metformin in Alzheimer’s disease: A snapshot. *Front. Pharmacol.* 12, 728315 (2021).
4. Weinstein, G. *et al.* Association of metformin, sulfonylurea and insulin use with brain structure and function and risk of dementia and Alzheimer’s disease: Pooled analysis from 5 cohorts. *PLoS One* 14, e0212293 (2019).
5. Royea, J., Zhang, L., Tong, X.-K. & Hamel, E. Angiotensin IV receptors mediate the cognitive and cerebrovascular benefits of losartan in a mouse model of Alzheimer’s disease. *J. Neurosci.* 37, 5562–5573 (2017).
6. Kehoe, P. G. *et al.* Safety and efficacy of losartan for the reduction of brain atrophy in clinically diagnosed Alzheimer’s disease (the RADAR trial): a double-blind, randomised, placebo-controlled, phase 2 trial. *Lancet Neurol.* 20, 895–906 (2021).
7. Miller, B. W., Willett, K. C. & Desilets, A. R. Rosiglitazone and pioglitazone for the treatment of Alzheimer’s disease. *Ann. Pharmacother.* 45, 1416–1424 (2011).
8. Galimberti, D. & Scarpini, E. Pioglitazone for the treatment of Alzheimer’s disease. *Expert Opin. Investig. Drugs* 26, 97–101 (2017).
9. Elkahloun, A. G., Hafko, R. & Saavedra, J. M. An integrative genome-wide transcriptome reveals that candesartan is neuroprotective and a candidate therapeutic for Alzheimer’s disease. *Alzheimers. Res. Ther.* 8, 5 (2016).
10. Torika, N., Asraf, K., Apte, R. N. & Fleisher-Berkovich, S. Candesartan ameliorates brain inflammation associated with Alzheimer’s disease. *CNS Neurosci. Ther.* 24, 231–242 (2018).
11. Kaeberlein, M. & Galvan, V. Rapamycin and Alzheimer’s disease: Time for a clinical trial? *Sci. Transl. Med.* 11, eaar4289 (2019).
12. Cai, Z. & Yan, L.-J. Rapamycin, autophagy, and Alzheimer’s disease. *J. Biochem. Pharmacol. Res.* 1, 84–90 (2013).
13. Tousi, B. The emerging role of bexarotene in the treatment of Alzheimer’s disease: current evidence. *Neuropsychiatr. Dis. Treat.* 11, 311–315 (2015).
14. Cummings, J. L. *et al.* Double-blind, placebo-controlled, proof-of-concept trial of bexarotene Xin moderate Alzheimer’s disease. *Alzheimers. Res. Ther.* 8, 4 (2016).
15. Matsunaga, S. *et al.* Lithium as a treatment for Alzheimer’s disease: A systematic review and meta-analysis. *J. Alzheimers. Dis.* 48, 403–410 (2015).
16. Forlenza, O. V., De-Paula, V. J. R. & Diniz, B. S. O. Neuroprotective effects of lithium: implications for the treatment of Alzheimer’s disease and related neurodegenerative disorders. *ACS Chem. Neurosci.* 5, 443–450 (2014).
17. Choi, Y. *et al.* Minocycline attenuates neuronal cell death and improves cognitive impairment in Alzheimer’s disease models. *Neuropsychopharmacology* 32, 2393–2404 (2007).
18. Budni, J. *et al.* The anti-inflammatory role of minocycline in Alzheimer´s disease. *Curr. Alzheimer Res.* 13, 1319–1329 (2016).
19. Turner, R. S. *et al.* Nilotinib effects on safety, tolerability, and biomarkers in Alzheimer’s disease. *Ann. Neurol.* 88, 183–194 (2020).
20. Lonskaya, I., Hebron, M. L., Selby, S. T., Turner, R. S. & Moussa, C. E.-H. Nilotinib and bosutinib modulate pre-plaque alterations of blood immune markers and neuro-inflammation in Alzheimer’s disease models. *Neuroscience* 304, 316–327 (2015).
21. Zhang, C. T. *et al.* Montelukast ameliorates streptozotocin-induced cognitive impairment and neurotoxicity in mice. *Neurotoxicology* 57, 214–222 (2016).
22. Lai, J. *et al.* Montelukast targeting the cysteinyl leukotriene receptor 1 ameliorates Aβ1-42-induced memory impairment and neuroinflammatory and apoptotic responses in mice. *Neuropharmacology* 79, 707–714 (2014).
23. Nygaard, H. B. *et al.* A phase Ib multiple ascending dose study of the safety, tolerability, and central nervous system availability of AZD0530 (saracatinib) in Alzheimer’s disease. *Alzheimers. Res. Ther.* 7, 35 (2015).
24. Kaufman, A. C. *et al.* Fyn inhibition rescues established memory and synapse loss in Alzheimer mice: Fyn Inhibition by AZD0530. *Ann. Neurol.* 77, 953–971 (2015).
25. Ma, J. *et al.* Fluoxetine attenuates the impairment of spatial learning ability and prevents neuron loss in middle-aged APPswe/PSEN1dE9 double transgenic Alzheimer’s disease mice. *Oncotarget* 8, 27676–27692 (2017).
26. Zhou, C.-N. *et al.* Fluoxetine delays the cognitive function decline and synaptic changes in a transgenic mouse model of early Alzheimer’s disease. *J. Comp. Neurol.* 527, 1378–1387 (2019).
27. Wang, J. *et al.* Carvedilol as a potential novel agent for the treatment of Alzheimer’s disease. *Neurobiol. Aging* 32, 2321.e1–12 (2011).
28. Liu, J. & Wang, M. Carvedilol protection against endogenous Aβ-induced neurotoxicity in N2a cells. *Cell Stress Chaperones* 23, 695–702 (2018).
29. Vallée, A., Vallée, J.-N., Guillevin, R. & Lecarpentier, Y. Riluzole: a therapeutic strategy in Alzheimer’s disease by targeting the WNT/β-catenin pathway. *Aging (Albany NY)* 12, 3095–3113 (2020).
30. Pereira, A. C. *et al.* Age and Alzheimer’s disease gene expression profiles reversed by the glutamate modulator riluzole. *Mol. Psychiatry* 22, 296–305 (2017).
31. Porsteinsson, A. P. *et al.* Effect of citalopram on agitation in Alzheimer disease: the CitAD randomized clinical trial: The CitAD randomized clinical trial. *JAMA* 311, 682–691 (2014).
32. Leonpacher, A. K. *et al.* Effects of citalopram on neuropsychiatric symptoms in Alzheimer’s dementia: Evidence from the CitAD study. *Am. J. Psychiatry* 173, 473–480 (2016).
33. Balducci, C. & Forloni, G. Doxycycline for Alzheimer’s disease: Fighting β-amyloid oligomers and neuroinflammation. *Front. Pharmacol.* 10, 738 (2019).
34. Molloy, D. W., Standish, T. I., Zhou, Q., Guyatt, G. & DARAD Study Group. A multicenter, blinded, randomized, factorial controlled trial of doxycycline and rifampin for treatment of Alzheimer’s disease: the DARAD trial: Doxycycline and rifampin for Alzheimer’s. *Int. J. Geriatr. Psychiatry* 28, 463–470 (2013).
35. Sparks, D. L. *et al.* Atorvastatin for the treatment of mild to moderate Alzheimer disease: preliminary results: Preliminary results. *Arch. Neurol.* 62, 753–757 (2005).
36. Feldman, H. H. *et al.* Randomized controlled trial of atorvastatin in mild to moderate Alzheimer disease: LEADe. *Neurology* 74, 956–964 (2010).
37. Folch, J. *et al.* Masitinib for the treatment of mild to moderate Alzheimer’s disease. *Expert Rev. Neurother.* 15, 587–596 (2015).
38. Piette, F. *et al.* Masitinib as an adjunct therapy for mild-to-moderate Alzheimer’s disease: a randomised, placebo-controlled phase 2 trial. *Alzheimers. Res. Ther.* 3, 16 (2011).
39. de Jong, D. L. K. *et al.* Effects of nilvadipine on cerebral blood flow in patients with Alzheimer disease: A randomized trial. *Hypertension* 74, 413–420 (2019).
40. Kennelly, S. P. *et al.* Demonstration of safety in Alzheimer’s patients for intervention with an anti-hypertensive drug Nilvadipine: results from a 6-week open label study. *Int. J. Geriatr. Psychiatry* 26, 1038–1045 (2011).
41. RePORTER. *Nih.gov* https://reporter.nih.gov/search/I615LI0fwkScy8VY5yYpcA/project-details/10185192.
42. Sanchez, P. E. *et al.* Levetiracetam suppresses neuronal network dysfunction and reverses synaptic and cognitive deficits in an Alzheimer’s disease model. *Proc. Natl. Acad. Sci. U. S. A.* 109, E2895-903 (2012).
43. Musaeus, C. S., Shafi, M. M., Santarnecchi, E., Herman, S. T. & Press, D. Z. Levetiracetam alters oscillatory connectivity in Alzheimer’s disease. *J. Alzheimers. Dis.* 58, 1065–1076 (2017).
44. Wang, H. *et al.* Pretreatment with antiasthmatic drug ibudilast ameliorates Aβ 1-42-induced memory impairment and neurotoxicity in mice. *Pharmacol. Biochem. Behav.* 124, 373–379 (2014).
45. Oliveros, G. *et al.* Multi-scale predictive modeling discovers Ibudilast as a polypharmacological agent to improve hippocampal-dependent spatial learning and memory and mitigate plaque and tangle pathology in a transgenic rat model of Alzheimer’s disease. *bioRxiv* (2021) doi:10.1101/2021.04.06.438662.
46. Oz, M., Lorke, D. E. & Petroianu, G. A. Methylene blue and Alzheimer’s disease. *Biochem. Pharmacol.* 78, 927–932 (2009).
47. Paban, V. *et al.* Therapeutic and preventive effects of methylene blue on Alzheimer’s disease pathology in a transgenic mouse model. *Neuropharmacology* 76 Pt A, 68–79 (2014).
48. Chau, S., Herrmann, N., Ruthirakuhan, M. T., Chen, J. J. & Lanctôt, K. L. Latrepirdine for Alzheimer’s disease. *Cochrane Database Syst. Rev.* CD009524 (2015) doi:10.1002/14651858.CD009524.pub2.
49. Bharadwaj, P. R. *et al.* Latrepirdine: molecular mechanisms underlying potential therapeutic roles in Alzheimer’s and other neurodegenerative diseases. *Transl. Psychiatry* 3, e332–e332 (2013).
50. Yao, Z.-G. *et al.* Valproate improves memory deficits in an Alzheimer’s disease mouse model: investigation of possible mechanisms of action. *Cell. Mol. Neurobiol.* 34, 805–812 (2014).
51. Hu, J.-P. *et al.* Valproate reduces tau phosphorylation via cyclin-dependent kinase 5 and glycogen synthase kinase 3 signaling pathways. *Brain Res. Bull.* 85, 194–200 (2011).
52. La, A. L. *et al.* Long-term trazodone use and cognition: A potential therapeutic role for slow-wave sleep enhancers. *J. Alzheimers. Dis.* 67, 911–921 (2019).
53. Ashford, J. W. Treatment of Alzheimer’s disease: Trazodone, sleep, serotonin, norepinephrine, and future directions. *J. Alzheimers. Dis.* 67, 923–930 (2019).
54. Bartlomé, P., King, K. S., Matsuo, F. & Wood, J. S. Cognitive decline with nortriptyline use in a patient with dementia of the Alzheimer’s type. *West. J. Med.* 156, 75–77 (1992).
55. Koźmiński, P., Halik, P. K., Chesori, R. & Gniazdowska, E. Overview of dual-acting drug methotrexate in different neurological diseases, autoimmune pathologies and cancers. *Int. J. Mol. Sci.* 21, 3483 (2020).
56. Newby, D. *et al.* Methotrexate and relative risk of dementia amongst patients with rheumatoid arthritis: a multi-national multi-database case-control study. *Alzheimers. Res. Ther.* 12, 38 (2020).
57. Prior, M., Dargusch, R., Ehren, J. L., Chiruta, C. & Schubert, D. The neurotrophic compound J147 reverses cognitive impairment in aged Alzheimer’s disease mice. *Alzheimers. Res. Ther.* 5, 25 (2013).
58. Prior, M. *et al.* Selecting for neurogenic potential as an alternative for Alzheimer’s disease drug discovery. *Alzheimers. Dement.* 12, 678–686 (2016).
59. Risner, M. E. *et al.* Efficacy of rosiglitazone in a genetically defined population with mild-to-moderate Alzheimer’s disease. *Pharmacogenomics J.* 6, 246–254 (2006).
60. Escribano, L. *et al.* Rosiglitazone rescues memory impairment in Alzheimer’s transgenic mice: mechanisms involving a reduced amyloid and tau pathology. *Neuropsychopharmacology* 35, 1593–1604 (2010).
61. Kume, K. *et al.* Effects of telmisartan on cognition and regional cerebral blood flow in hypertensive patients with Alzheimer’s disease: Telmisartan on cognition and rCBF in AD. *Geriatr. Gerontol. Int.* 12, 207–214 (2012).
62. Singh, B., Sharma, B., Jaggi, A. S. & Singh, N. Attenuating effect of lisinopril and telmisartan in intracerebroventricular streptozotocin induced experimental dementia of Alzheimer’s disease type: possible involvement of PPAR-γ agonistic property. *J. Renin Angiotensin Aldosterone Syst.* 14, 124–136 (2013).
63. Zhou, B. *et al.* Association between exenatide use and incidence of Alzheimer’s disease. *Alzheimers Dement. (N. Y.)* 7, e12139 (2021).
64. An, J. *et al.* Exenatide alleviates mitochondrial dysfunction and cognitive impairment in the 5×FAD mouse model of Alzheimer’s disease. *Behav. Brain Res.* 370, 111932 (2019).
65. Butchart, J. *et al.* Etanercept in Alzheimer disease: A randomized, placebo-controlled, double-blind, phase 2 trial. *Neurology* 84, 2161–2168 (2015).
66. Torres-Acosta, N., O’Keefe, J. H., O’Keefe, E. L., Isaacson, R. & Small, G. Therapeutic potential of TNF-α inhibition for Alzheimer’s disease prevention. *J. Alzheimers. Dis.* 78, 619–626 (2020).
67. Cummings, J. L. *et al.* Effect of dextromethorphan-quinidine on agitation in patients with Alzheimer disease dementia: A randomized clinical trial: A randomized clinical trial. *JAMA* 314, 1242–1254 (2015).
68. Ballard, C., Sharp, S. & Corbett, A. Dextromethorphan and quinidine for treating agitation in patients with Alzheimer disease dementia. *JAMA* 314, 1233–1235 (2015).
69. Zhang, X.-Z., Li, X.-J. & Zhang, H.-Y. Valproic acid as a promising agent to combat Alzheimer’s disease. *Brain Res. Bull.* 81, 3–6 (2010).
70. Xuan, A.-G. *et al.* Valproic acid alleviates memory deficits and attenuates amyloid-β deposition in transgenic mouse model of Alzheimer’s disease. *Mol. Neurobiol.* 51, 300–312 (2015).
71. Wang, J. *et al.* Valsartan lowers brain beta-amyloid protein levels and improves spatial learning in a mouse model of Alzheimer disease. *J. Clin. Invest.* 117, 3393–3402 (2007).
72. Yang, W.-N. *et al.* The effects of valsartan on cognitive deficits induced by aluminum trichloride and d-galactose in mice. *Neurol. Res.* 36, 651–658 (2014).
73. Fremont, R. *et al.* Tolcapone treatment for cognitive and behavioral symptoms in behavioral variant frontotemporal dementia: A placebo-controlled crossover study. *J. Alzheimers. Dis.* 75, 1391–1403 (2020).
74. Grossmann, K. Anticoagulants for treatment of Alzheimer’s disease. *J. Alzheimers. Dis.* 77, 1373–1382 (2020).
75. Wang, J. *et al.* Investigation of nebivolol as a novel therapeutic agent for the treatment of Alzheimer’s disease. *J. Alzheimers. Dis.* 33, 1147–1156 (2013).
76. Sanders, O. Sildenafil for the treatment of Alzheimer’s disease: A systematic review. *J. Alzheimers Dis. Rep.* 4, 91–106 (2020).
77. Chumakov, I. *et al.* Combining two repurposed drugs as a promising approach for Alzheimer’s disease therapy. *Sci. Rep.* 5, 7608 (2015).
78. in t’ Veld, B. A. *et al.* Nonsteroidal antiinflammatory drugs and the risk of Alzheimer’s disease. *N. Engl. J. Med.* 345, 1515–1521 (2001).
79. Aisen, P. S. Inflammation and Alzheimer’s disease: mechanisms and therapeutic strategies. *Gerontology* 43, 143–149 (1997).
80. Cao, S., Yu, L., Mao, J., Wang, Q. & Ruan, J. Uncovering the molecular mechanism of actions between pharmaceuticals and proteins on the AD network. *PLoS One* 10, e0144387 (2015).
81. O’Neill, E., Kwok, B., Day, J. S., Connor, T. J. & Harkin, A. Amitriptyline protects against TNF-α-induced atrophy and reduction in synaptic markers via a Trk-dependent mechanism. *Pharmacol. Res. Perspect.* 4, e00195 (2016).
82. Yang, L., Liu, Y., Wang, Y., Li, J. & Liu, N. Azeliragon ameliorates Alzheimer’s disease via the Janus tyrosine kinase and signal transducer and activator of transcription signaling pathway. *Clinics (Sao Paulo)* 76, e2348 (2021).
83. Pilipenko, V. *et al.* Very low doses of muscimol and baclofen ameliorate cognitive deficits and regulate protein expression in the brain of a rat model of streptozocin-induced Alzheimer’s disease. *Eur. J. Pharmacol.* 818, 381–399 (2018).
84. Watt, G. & Karl, T. In vivo evidence for therapeutic properties of cannabidiol (CBD) for Alzheimer’s disease. *Front. Pharmacol.* 8, 20 (2017).
85. Breitner, J. C. *et al.* Delayed onset of Alzheimer’s disease with nonsteroidal anti-inflammatory and histamine H2 blocking drugs. *Neurobiol. Aging* 16, 523–530 (1995).
86. Li, Z.-Y. *et al.* Clemastine attenuates AD-like pathology in an AD model mouse via enhancing mTOR-mediated autophagy. *Exp. Neurol.* 342, 113742 (2021).
87. Beeri, M. S. *et al.* Corticosteroids, but not NSAIDs, are associated with less Alzheimer neuropathology. *Neurobiol. Aging* 33, 1258–1264 (2012).
88. Moura, F. C. D. *et al.* Behavioral, neurochemical and histological changes in the use of low doses of Naltrexone and Donepezil in the treatment in experimental model of Alzheimer’s disease by induction of β-Amyloid1-42 in rats. *World Sci. Res.* 6, 5–13 (2019).
89. Chen, S., Liu, A.-R., An, F.-M., Yao, W.-B. & Gao, X.-D. Amelioration of neurodegenerative changes in cellular and rat models of diabetes-related Alzheimer’s disease by exendin-4. *Age (Dordr.)* 34, 1211–1224 (2012).
90. Irwin, R. W., Solinsky, C. M. & Brinton, R. D. Frontiers in therapeutic development of allopregnanolone for Alzheimer’s disease and other neurological disorders. *Front. Cell. Neurosci.* 8, 203 (2014).
91. Aisen, P. S., Marin, D. B., Brickman, A. M., Santoro, J. & Fusco, M. Pilot tolerability studies of hydroxychloroquine and colchicine in Alzheimer disease. *Alzheimer Dis. Assoc. Disord.* 15, 96–101 (2001).
92. Wilkinson, B. L. *et al.* Ibuprofen attenuates oxidative damage through NOX2 inhibition in Alzheimer’s disease. *Neurobiol. Aging* 33, 197.e21–32 (2012).
93. Freiherr, J. *et al.* Intranasal insulin as a treatment for Alzheimer’s disease: a review of basic research and clinical evidence. *CNS Drugs* 27, 505–514 (2013).
